# Effect of Gene-based vs. Standard Weight-Loss Recommendations on Anthropometry, Lipid and Glucose Markers, and Dietary Intake: The MyGeneMyDiet® Study

**DOI:** 10.1101/2025.03.02.25323063

**Authors:** Jacus S. Nacis, Jason Paolo H. Labrador, Diana Glades D. Ronquillo, John Chris Rementilla, Marietta P. Rodriguez, Marilou L. Madrid, Ruby D. Frane, Noelle Lyn C. Santos, Aurora Maria Francesca D. Dablo, Julianne Janine V. Carrillo, Mikko Glen Fernandez, Diane Jane V. Fanio, Debbie Ann S. Martirez, Malaya V. Paller, Hannah Sancha S. Monje, Rachelle Mae V. Cabigan, Angela Alexis Bausas, Gyra Marie Agra, Reminda C. Bunhiyan, Fränzel J.B. van Duijnhoven, Gerard Bryan Gonzales

## Abstract

**Background:** Gene-based nutrition recommendations have emerged as a strategy for weight management, but their effectiveness over standard advice remains uncertain.

**Objective:** This study evaluated MyGeneMyDiet® recommendations versus standard advice on anthropometry, biochemical markers, and dietary intake in overweight and obese Filipino adults over 12 months.

**Methods:** In this 12-month randomized controlled trial, participants received either MyGeneMyDiet® or standard advice. Both groups underwent regular nutrition counseling during the active phase (months 0–6) before transitioning to free-living conditions (months 6–12). Primary outcomes included weight, BMI, waist circumference, and body fat percentage; secondary outcomes were dietary intake and biochemical markers. Analyses followed an intention-to-treat approach, with paired t-tests for within-group comparisons and ANCOVA for between-group differences. Sensitivity analyses used Last Observation Carried Forward (LOCF) and Inverse Probability of Attrition Weighting (IPAW) to address loss-to-follow-up.

**Results:** Of 136 screened, 52 participants (19–59 years) were enrolled (MyGeneMyDiet®, n = 29; standard recommendation, n = 23), with 27 completing the study (MyGeneMyDiet®, n = 15; standard recommendation, n = 12). Weight changes over 12 months were minimal, with no substantial differences between groups. At month 6, baseline-adjusted analyses showed no meaningful differences in weight (−0.36 kg [95% CI: −1.77, 1.04]), BMI (0.11 kg/m² [−0.51, 0.73]), waist circumference (−0.27 cm [−2.23, 1.69]), or body fat percentage (0.92% [−0.86, 1.05]). These trends persisted on month 12. While both groups reduced dietary intake, the MyGeneMyDiet® group showed greater decreases in total calories (−461 kcal, P = 0.001), protein (−12 g, P = 0.007), carbohydrates (−46 g, P = 0.015), and fat (−22 g, P = 0.014), though between-group differences remained modest.

**Conclusions:** Gene-based and standard weight management advice led to comparable weight and metabolic outcomes over 12 months. While gene-based recommendations influenced dietary intake, these changes did not improve anthropometric or biochemical outcomes.

This trial was registered at clinicaltrials.gov as NCT05098899.

## Introduction

Genetic factors have been shown to contribute to people’s weight and inter-individual variability in response to weight loss [1–3]. Variants in genes such as the fat mass and obesity-associated (*FTO*) gene have been implicated in obesity-related phenotypes through their influence on appetite regulation and energy balance [4–5]. Notably, the *FTO* rs9939609 polymorphism has been extensively studied, demonstrating that carriers of the risk A allele, particularly those with low physical activity levels, face a higher susceptibility to obesity [7–9]. However, evidence suggests that being physically active can attenuate this genetic risk [8, 10].

Similarly, polymorphisms in the uncoupling protein 1 (*UCP1*) gene, such as rs1800592, have been associated with altered energy expenditure and thermogenesis, potentially leading to resistance to weight loss when following energy-restricted diets [11–15]. Additionally, the transcription factor 7-like 2 (*TCF7L2*) gene variant rs7903146, strongly linked to type 2 diabetes, is also associated with an elevated risk of obesity and related metabolic phenotypes, including impaired glucose regulation and dyslipidemia [16–21]. Evidence suggests that dietary intake, particularly saturated fat, may amplify metabolic risks in individuals carrying this variant, highlighting the potential for tailored dietary recommendations to mitigate such risks [18].

While prior research has examined the interaction between genetic risk and dietary intake [22, 23], the efficacy of gene-based nutritional recommendations in improving weight-loss outcomes remains inconclusive. Studies have reported mixed findings, with some observing no added benefit over standard recommendations [23, 24], while others suggest significant improvements in weight loss, dietary behaviors, or lifestyle modifications with personalized advice based on genetic profiles [25–28]. Notably, awareness of genetic predisposition alone does not consistently enhance weight management outcomes [22, 23, 27, 28].

The generalizability of gene-diet interactions remains limited due to small sample sizes, heterogeneity in intervention designs, and inconsistent adherence reporting [29–31]. Additionally, prior studies often focus on short-term outcomes [32, 33], leaving the long-term sustainability of gene-based recommendations largely unexplored. Evidence comparing the effectiveness of gene-based versus standard recommendations in real-world settings is scarce, particularly in interventions that assess both immediate and follow-up phases.

Moreover, while genetic-based recommendations have shown potential to influence dietary behavior, few studies have comprehensively assessed their impact on broader dietary patterns alongside anthropometric and biochemical outcomes [34–36]. This leaves an important question unanswered: Do gene-based dietary recommendations lead to measurable health benefits over time?

This study aims to address these gaps by evaluating the effects of the MyGeneMyDiet® recommendation, which incorporates genetic risk information into dietary and physical activity recommendations, on anthropometric, biochemical, and dietary outcomes over 12 months. The two-phase design of this randomized controlled trial enables the assessment of both active intervention and post-intervention phases, to provide insights into the sustainability of behavior changes once structured support is withdrawn.

## Methods

### Study design

This was a 12-month, parallel-group, single-blinded, randomized controlled trial designed to evaluate the effect of gene-based nutrition recommendations on weight loss, blood biomarkers, and dietary outcomes in overweight and obese Filipino adults (clinicaltrials.gov identifier: NCT05098899). Study outcomes were assessed at five time points: baseline (month 0), and months 3, 6, 9, and 12 and entailed an active (month 0-6) and inactive (month 6-12) phases of intervention. All participants received regular sessions of nutrition counseling and monitoring of outcomes during the active phase, while only follow-up data collection without nutrition counseling was conducted during the inactive phase.

The trial was conducted in accordance with the Declaration of Helsinki and was approved by the Food and Nutrition Research Institute (FNRI) Institutional Ethics Review Committee (FIERC-2021-001). Written informed consent was obtained from all study participants. A detailed study design and intervention protocol have been published elsewhere [37] and briefly described here.

### Study participants

Participants were recruited primarily from Metro Manila and nearby provinces between December 2021 to November 2022, through invitation letters, social media promotional materials, and snowball sampling. Orientation sessions were conducted via videoconferencing platforms, and FIERC-approved consent forms were emailed to the potentially eligible participants after the orientation.

Eligible individuals were overweight or obese adults aged 19 to 59 years, with a BMI between 25 and 40 kg/m^2^, and carriers of at least one risk allele: A for *FTO* rs9939609, C for *TCF7L2* rs7903146, and G for *UCP1* rs1800592. Supplementary Table 1 summarizes the detailed eligibility criteria.

### Screening and data collection protocols

Owing to the COVID-19 pandemic during the screening of participants, all in-person screening and data collection procedures were conducted at the FNRI, Taguig City, Metro Manila, while adhering to national and local COVID-19 guidelines. Prior to each session, the participants and research team members completed an online health declaration form, with those recently exposed to COVID-19 cases required to follow prescribed isolation protocols. On the day of data collection, all individuals underwent mandatory on-site temperature checks, hand sanitation, and rapid antigen COVID-19 testing. The data collection process was organized into five stations: COVID-19 testing, registration, anthropometric measurements, blood collection, and physician consultation. To minimize contact, participants spent no more than 15 minutes at each station and adhered to strict queuing protocols. After each session, both participants and research team members were monitored for COVID-19 symptoms over a 14-day period. These protocols were consistently applied during outcomes assessment at months 0, 3, 6, 9, and 12, with modifications as necessary based on Metro Manila’s quarantine classifications.

### Genetic counseling sessions

All participants underwent genetic counseling before randomization into either the intervention or control arm. This process included pre- and post-genetic test counseling sessions, conducted virtually in a two-part, interactive format.

The pre-genetic test counseling occurred a few days after DNA sample collection and before the genotyping of samples. During this session, a genetic counselor provided education on genetics, covering topics such as genes, chromosomes, genetic mutations, environmental and genetic interactions, and procedures for genetic testing. The counselor also addressed the potential emotional and psychological impacts of genetic test results.

The post-genetic test counseling was scheduled for two weeks after the pre-genetic test session, coinciding with the release of genetic testing results. In this session, the genetic counselor disclosed the genotyping results and offered psychosocial support to help participants process and adapt to the new information.

### Randomization

Participants were stratified by BMI according to World Health Organization (WHO) cutoffs: overweight (BMI 25–30 kg/m²) and obese (BMI >30 kg/m²). To ensure balanced allocation, participants were randomly assigned to either the MyGeneMyDiet® group or the standard recommendation group using block randomization with block sizes of 4, 6, and 8. Random allocation was performed using Random Allocation Software by two research staff to maintain consistency and minimize bias.

The randomization involved preparing 66 identical, sequentially numbered, opaque sealed envelopes (SNOSE), with 33 envelopes designated for each treatment group. The sequential numbering was synchronized with participant registration and used to assign study numbers. Allocation was concealed from participants and researchers involved in the month 0 visit.

### Intervention strategy

The study employed a two-arm design consisting of an intervention arm and a control arm. Participants in the intervention arm received the MyGeneMyDiet® recommendations, a personalized, gene-based nutrition and lifestyle intervention tailored to the participants’ genetic profiles. These recommendations were developed by integrating information from participants’ genotyping results, anthropometric data such as BMI, waist circumference, and body fat percentage, biochemical test results, dietary intake, and physical activity levels.

The MyGeneMyDiet® focused on three specific genetic polymorphisms that have been associated with weight management and metabolic outcomes. These included *FTO* rs9939609, a variant related to weight, BMI, and physical activity; *UCP1* rs1800592, linked to calorie requirements; and *TCF7L2* rs7903146, associated with dietary fat intake. For each of these polymorphisms, participants received specific recommendations tailored to their genetic profiles.

For participants carrying the A risk allele of *FTO* rs9939609, the advice was to engage in 30 to 60 minutes of moderate-intensity aerobic physical activity daily. Those with the G risk allele of *UCP1* rs1800592 were advised to reduce their daily caloric intake by 150 kilocalories from the recommended daily calorie intake. Lastly, participants with the C risk allele of TCF7L2 rs7903146 were recommended the limit total caloric intake from fat to 15-20%, which falls within the lower range of the standard guideline of 15-30%.

Participants with multiple risk alleles received combined advice relevant to each polymorphism in their genetic profile. These recommendations were generated using decision trees and coded messages that reflected the interaction of genetic polymorphisms, clinical data, and lifestyle factors. The intervention was developed with the input of a Scientific Advisory Board that included experts in nutrition, genetics, and lifestyle medicine (Supplementary Material: Development of MyGeneMyDiet® Recommendations). Throughout the study, the intervention was delivered by registered nutritionist-dietitians (RNDs) through online counseling sessions at months 0, 3, 6, and 12. During these sessions, participants’ compliance was monitored, and their caloric and weight-loss goals were adjusted when necessary.

In the control arm, participants were provided with standard weight management advice, which was based on existing guidelines such as the Philippine Nutrition Practice Guidelines for the Screening and Management of Obesity, the 2012 Nutritional Guidelines for Filipinos, the *Pinggang Pinoy* (food plate) dietary model, and the 2020 WHO Guidelines on Physical Activity. These recommendations were derived from anthropometric and biochemical data, dietary intake, and physical activity levels, but did not consider participants’ genetic profiles. Like the intervention group, the control group also received online counseling in months 0, 3, 6, and 12. After the 12-month study was completed, participants in the control arm were provided with the MyGeneMyDiet® recommendations as part of their after-trial care.

### Outcomes assessment

The primary outcomes are differences in weight, BMI, waist circumference, and body fat percentage between the arms at both the active (month 6) and inactive phases (month 12). Secondary outcomes include differences in blood lipid profile, glycated hemoglobin, and dietary intake.

#### Anthropometric Measurements

Body weight and body fat percentage were determined using a body composition analyzer (Tanita-MC78U, Tanita Corporation, Japan). Height was measured with a stadiometer (Seca 217, Seca, Germany), and waist circumference obtained using a non-stretchable tape measure (Seca 203, Seca, Germany). All anthropometric measurements were taken twice, following standard procedures. A third measurement was taken if the difference between the two initial readings exceeded 0.3 kg for weight, 0.5 cm for height, or 0.5 cm for waist circumference. The average of all the reading was recorded as the final value for each measurement. BMI was calculated by dividing the weight into kilograms by the height in meters squared. Body fat percentage was classified using the chart developed by Gallagher et al. [38].

#### Dietary intake

Participants completed a three-day food diary for each month to estimate their mean energy and macronutrient intake throughout the trial. The food records included two non-consecutive weekdays and one weekend day. The proper method for recording food and beverage intake, including estimating portion sizes, was explained to participants in a separate orientation session. Each diary recorded all food and beverages consumed, providing details such as type, variety, brand, portion sizes, meal timing, preparation methods, and source of food (e.g., home-cooked or takeaway). Printed or electronic versions of the food diary were distributed prior to the follow-up sessions. RNDs checked and verified the diaries for completeness and accuracy two weeks before each online counseling session. Dietary analysis was conducted using the Philippine Food Composition Tables (PhilFCT), supplemented with international FCTs for items not listed in the local FCT.

#### Biochemical data and physical activity

Blood samples were collected at baseline and at months 3, 6, 9, and 12. Participants were instructed to fast for 10–12 hours before each specimen collection. Approximately 5 mL of venous blood was drawn by a trained phlebotomist and processed for the analysis of glycated hemoglobin (HbA1c) and lipid profiles.

Biochemical analyses were conducted by a third-party laboratory using ion-exchange high-performance liquid chromatography (HPLC) for HbA1c and colorimetric methods for lipid profiles. Quality control procedures were performed daily, including the use of low- and high-control samples. All quality control analyses were required to meet predefined standards before the processing of participant samples commenced.

Physical activity was assessed using the International Physical Activity Questionnaire – Short Form (IPAQ-SF), which included seven questions to evaluate the frequency and type of physical activity participants engaged in as part of their daily routines.

### Adverse event monitoring

All reported adverse events were evaluated by the principal investigator and the research physician to assess the severity and any potential relation to the study protocol. The study team ensured that any reportable events were communicated to the institutional ethics review committee (FIERC). Throughout the study, no severe adverse events were identified.

### Statistical analysis

Data were analyzed following the intention-to-treat (ITT) principle, ensuring that all participants originally randomized to either the standard recommendation or MyGeneMyDiet® groups were included in the final analysis. Descriptive statistics are presented as mean ± standard deviation (SD) for continuous variables and numbers with frequencies for categorical variables.

The normality of data distribution was assessed using the Shapiro-Wilk test, while Levene’s test evaluated homogeneity of variances between groups. To account for baseline differences, an Analysis of Covariance (ANCOVA) was performed, with baseline measurements included as covariates to control for their potential influence on outcomes. Within-group differences were assessed using paired t-tests.

To address missing data, the Last Observation Carried Forward (LOCF) method was used alongside Inverse Probability of Attrition Weighting (IPAW) as sensitivity analyses to assess potential biases from dropout. IPAW weights were calculated based on baseline characteristics, including age, gender, job classification, and weight. All statistical analyses were conducted using SPSS version 26 and R version 4.2.3. Statistical significance was set at P < 0.05.

## Results

### Study Participants

A total of 136 potential participants were screened for the study. Following genetic testing, 66 were randomized into the MyGeneMyDiet® (n = 33) and standard recommendation (n = 33) groups. Of these, 52 entered the trial at month 0 (MyGeneMyDiet®, n = 29; standard recommendation, n = 23), as 14 did not attend the baseline measurement session. During the active intervention phase (month 0–6), 49 participants remained (MyGeneMyDiet®, n = 26; standard recommendation, n = 23), while 27 continued through the inactive phase (month 6–12) (MyGeneMyDiet®, n = 15; standard recommendation, n = 12). The CONSORT diagram (Figure 1) illustrates the randomization process, as well as participant withdrawals and losses to follow-up.

**Figure 1:**
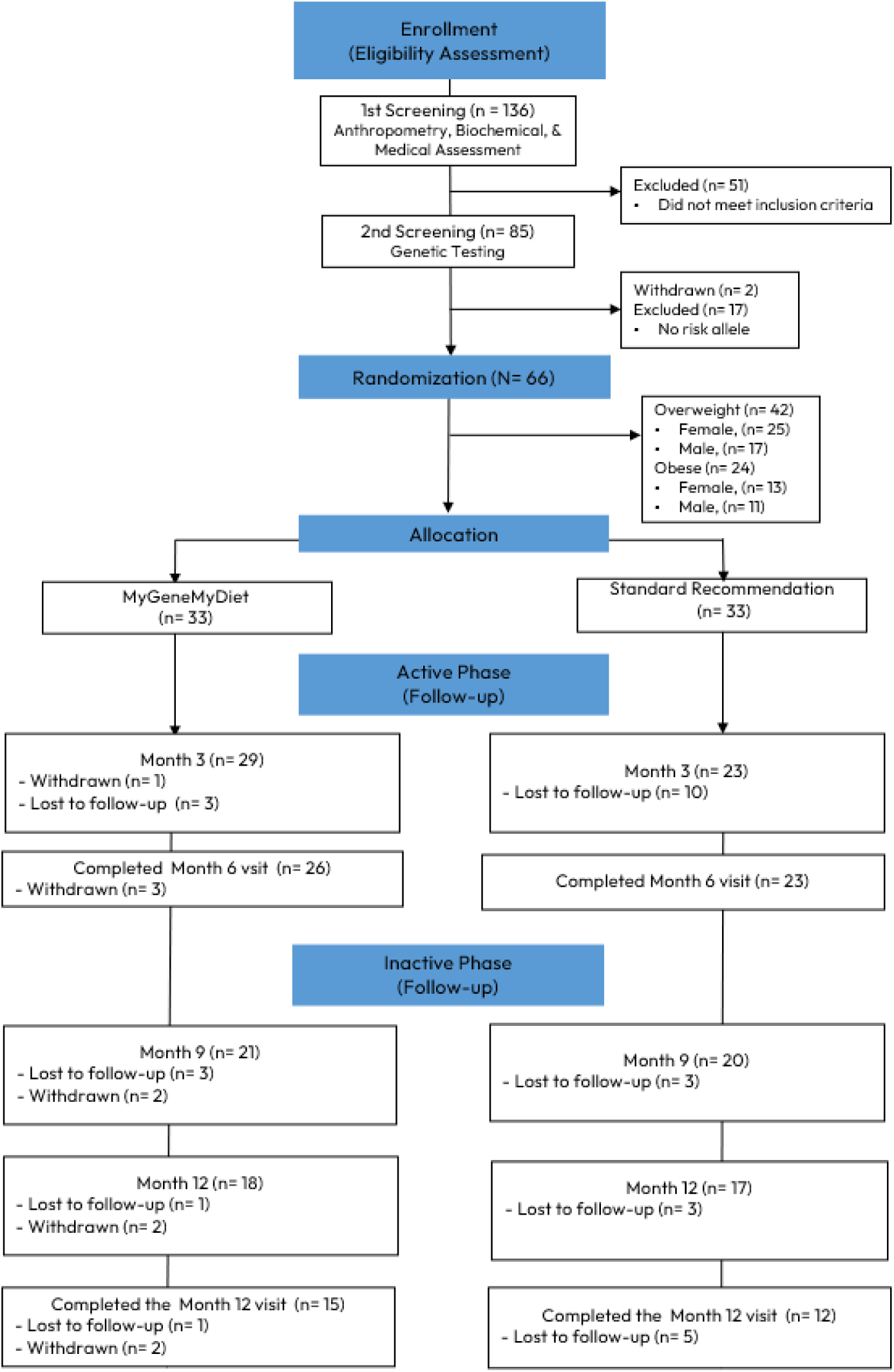
The CONSORT diagram of the study.

Baseline characteristics of the study participants are summarized in Table 1. Among the 52 participants, the majority were female (57.7%; n = 30), with most participants aged 18–29 years (44.2%; n = 23). Anthropometric measures were generally comparable between the two groups. The standard recommendation group had a mean (SD) weight of 76.6 (14.2) kg, BMI of 29.0 (2.9) kg/m², and waist circumference of 90.2 (9.2) cm, while the MyGeneMyDiet® group had a mean (SD) weight of 76.8 (14.2) kg, BMI of 29.9 (4.6) kg/m², and waist circumference of 91.0 (10.9) cm. However, the participants randomized to the MyGeneMyDiet® group had higher baseline mean body fat percentage (38.3 [7.7] %) than those in the standard recommendation group (33.8 [7.6] %). Baseline biochemical markers, including HbA1c, total cholesterol, HDL-cholesterol, LDL-cholesterol, and triglycerides, were similar between groups. Dietary intake showed slight variation, with the standard recommendation group reporting higher mean calorie, protein, carbohydrate, and fat intakes than the MyGeneMyDiet® group.

**Table 1.**
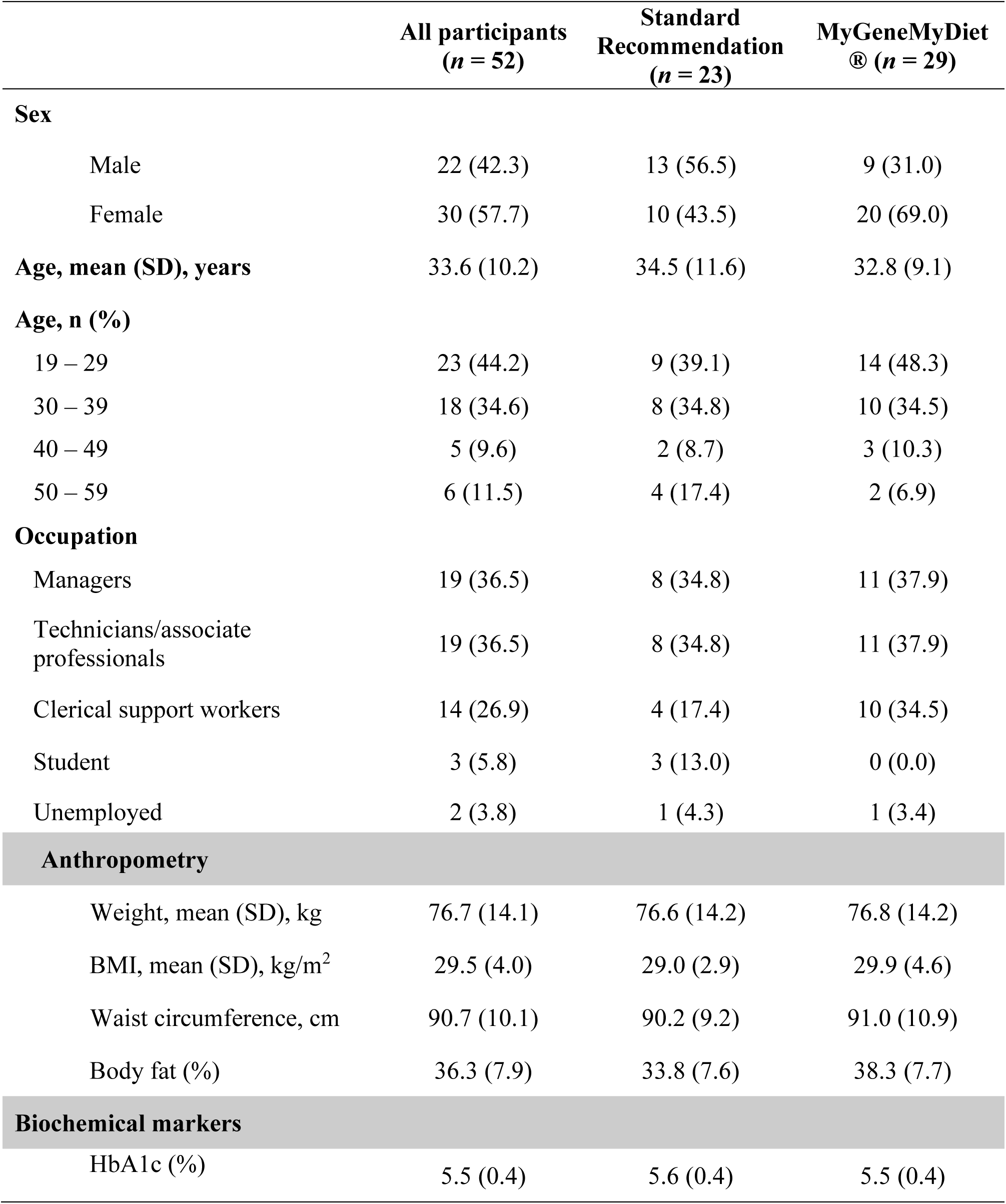
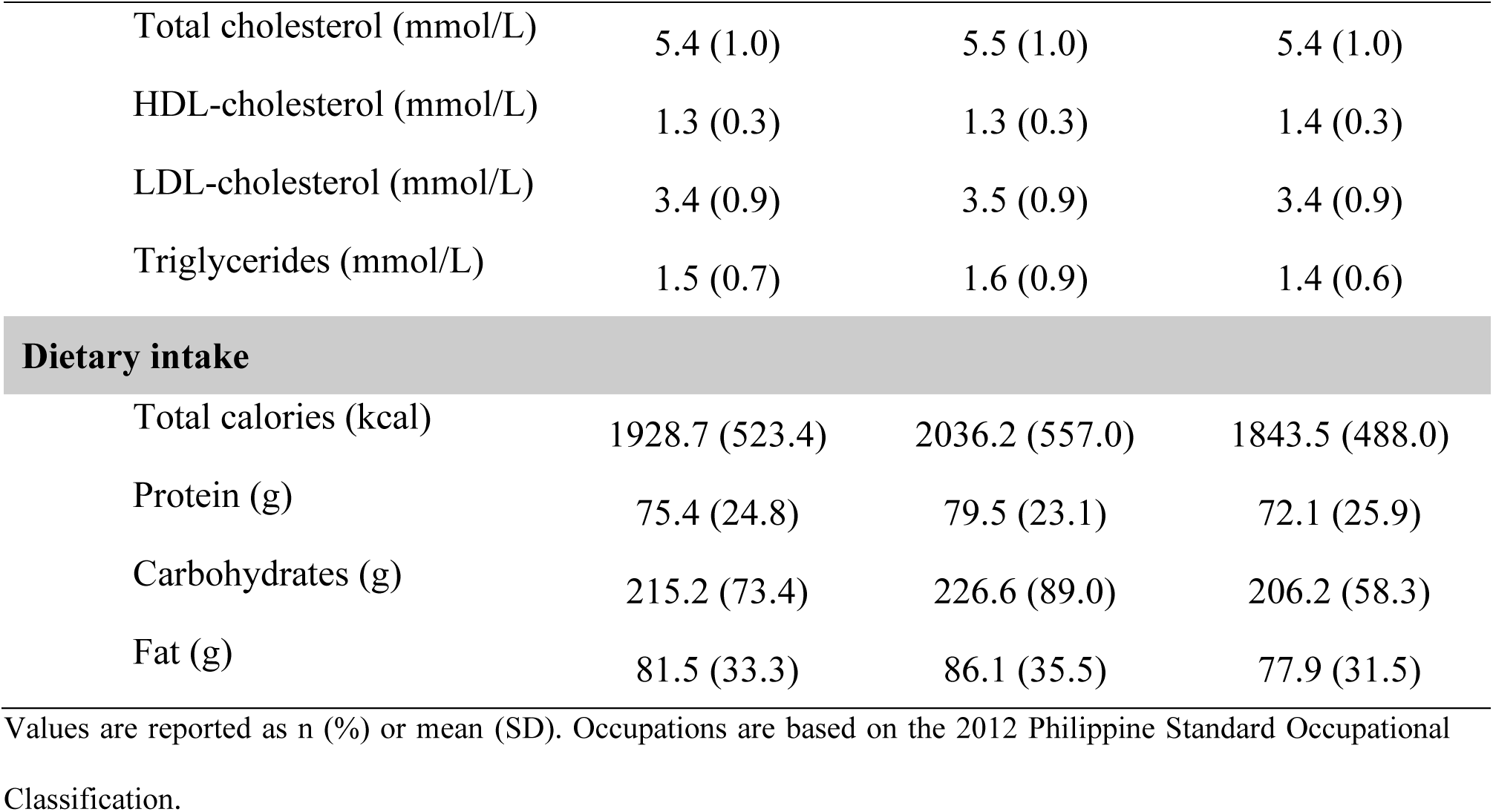
Participant characteristics at randomization.

In terms of genotype distribution, the AA variant of *FTO* rs9939609 was rare, observed only in the MyGeneMyDiet® group (n = 3, 10.3%) (Supplementary Table 2). Meanwhile, the CC genotype of *TCF7L2* rs7903146 was predominant in both groups (MyGeneMyDiet®: n = 29, 100%; standard recommendation: n = 21, 91.3%). For *UCP1* rs1800592, the GA genotype was the most common, appearing in 18 out of 29 participants (62.1%) in the MyGeneMyDiet® group and 12 out of 23 (52.2%) in the standard recommendation group.

### Engagement with the intervention of the study participants

Adherence to the MyGeneMyDiet® and standard weight-loss recommendations were evaluated at 3-, 6-, 9-, and 12-months using participants’ food diaries and responses to IPAQ-SF questionnaires collected during counseling sessions (Supplementary Table 3). We found no evidence to suggest differences in adherence between the groups at any time point. However, adherence was generally low, with rates mostly below 50% for both arms. This was circumvented by using loss-to-follow-up strategies such as LOCF and IPAW for the analysis.

### Weight Reduction Across Timepoints

Both groups exhibited a modest decrease in body weight over 12 months (Figure 2). The MyGeneMyDiet® group decreased from 76.7 kg at baseline to 71.7 kg at month 12, while the standard recommendation group showed a comparable reduction (76.6 kg to 71.7 kg).

**Figure 2:**
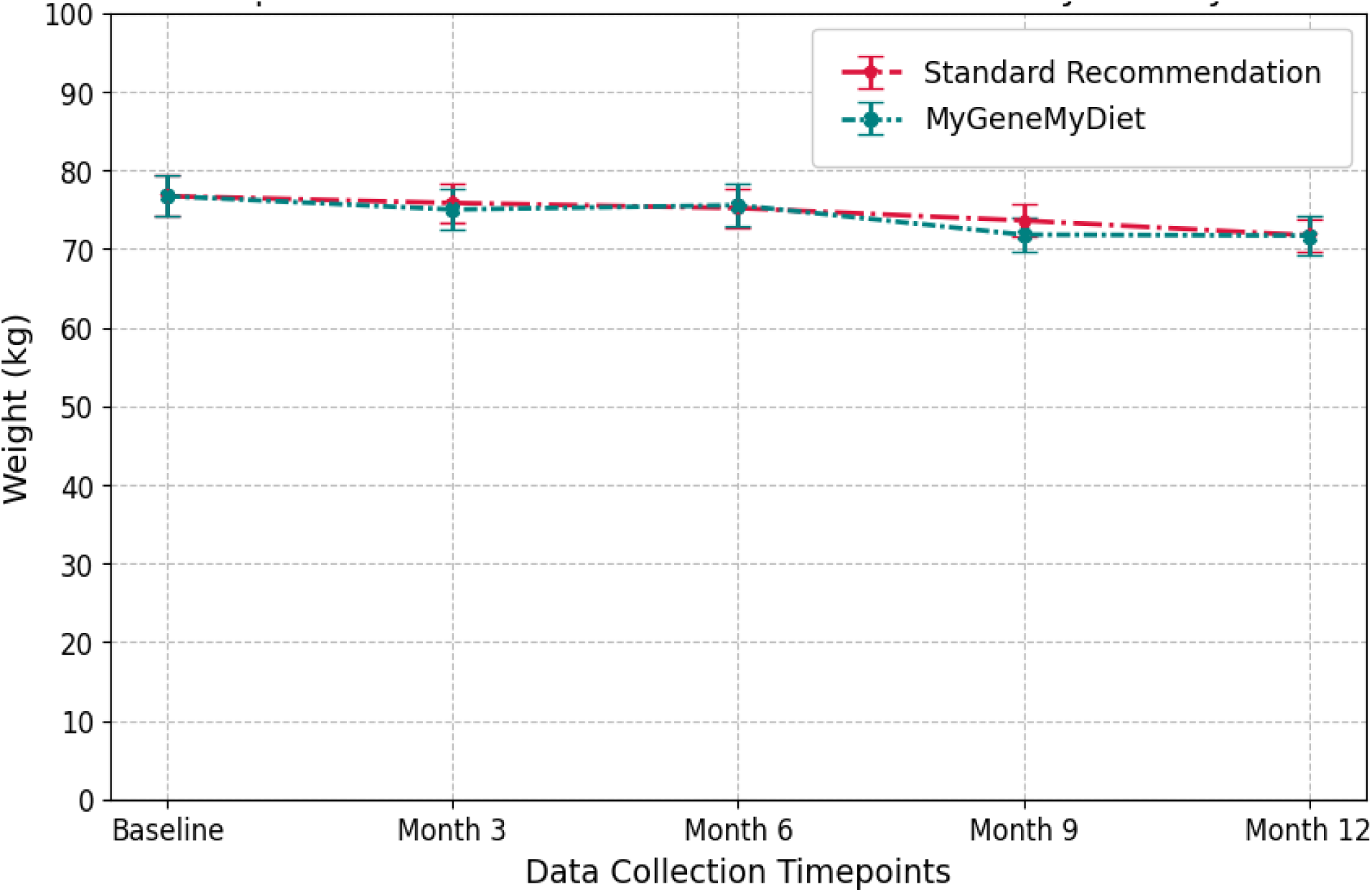
Changes in body weight (kg) over 12 months in the MyGeneMyDiet® and standard recommendation groups. Data are presented as means ± SEM. No significant differences were observed between groups at any time point (P > 0.05).

### Comparison of Outcomes Between Intervention Groups at Month 6 (Active Phase)

At month 6 (end of the active phase), there were no meaningful differences between the MyGeneMyDiet® and standard recommendation groups for most measured outcomes (Table 2). Anthropometric measures, including weight (difference: −0.36 kg [95% CI: −1.77, 1.04], P = 0.604), BMI (difference: 0.11 kg/m² [−0.51, 0.73], P = 0.716), waist circumference (difference: −0.27 cm [−2.23, 1.69], P = 0.781), and body fat percentage (difference: 0.92% [−0.86, 1.05], P = 0.848), were similar in the two groups.

**Table 2.** Differences in outcomes at the end of the active (month 6) and inactive phases (month 12) of the intervention.

| **ACTIVE PHASE (Month 6)** |  |  |  |  |
| --- | --- | --- | --- | --- |
|  | **Standard Recommendation**<br>**(*n* = 23)** | **MyGeneMyDiet®**<br>**(*n* = 29)** | **Difference [95% CI]** | ***P* – value\*** |
| **Anthropometry** |  |  |  |  |
| Weight (kg) | 75.17 | 75.54 | -0.36 [-1.77, 1.04] | 0.604 |
| BMI (kg/m<sup>2</sup>) | 29.22 | 29.11 | 0.11 [-0.51, 0.73] | 0.716 |
| Waist circumference (cm) | 88.61 | 88.88 | -0.27 [-2.23, 1.69] | 0.781 |
| Body fat (%) | 36.07 | 35.98 | 0.92 [-0.86, 1.05] | 0.848 |
| **Biochemical markers** |  |  |  |  |
| HbA1c (%) | 5.60 | 5.49 | 0.11 [0.01, 0.20] | 0.038 |
| Total cholesterol (mmol/L) | 5.18 | 5.30 | -0.12 [-0.57, 0.32] | 0.222 |
| HDL-cholesterol (mmol/L) | 1.34 | 1.27 | 0.07 [-0.05, 0.18] | 0.241 |
| LDL-cholesterol (mmol/L) | 3.18 | 3.23 | -0.05 [-0.48, 0.39] | 0.826 |
| Triglycerides (mmol/L) | 1.48 | 1.76 | -0.28 [-0.99, 0.42] | 0.420 |
| **Dietary intake** |  |  |  |  |
| Total calories (kcal) | 1667.5 | 1524.4 | 143.1 [-138.50, 424.71] | 0.312 |
| Protein (g) | 65.73 | 62.39 | 3.34 [-7.80, 14.48] | 0.549 |
| Carbohydrates (g) | 194.93 | 171.31 | 23.62 [-9.98, 57.23] | 0.164 |
| Fat (g) | 67.50 | 66.56 | 0.94 [-14.96, 16.85] | 0.905 |

INACTIVE PHASE (Month 12)
|  | Standard Recommendation<br>(*n* = 12) | MyGeneMyDiet®<br>(*n* = 15) | Difference [95% CI] | *P* – value* |
| --- | --- | --- | --- | --- |
| **Anthropometry** |  |  |  |  |
| Weight (kg) | 71.71 | 71.67 | 0.04 [-3.41, 3.49] | 0.981 |
| BMI (kg/m<sup>2</sup>) | 28.03 | 27.88 | 0.15 [-1.24, 1.52] | 0.831 |
| Waist circumference (cm) | 84.45 | 84.57 | -0.12 [-4.58, 4.33] | 0.955 |
| Body fat (%) | 34.87 | 34.60 | 0.27 [-2.02, 2.56] | 0.809 |
| **Biochemical markers** |  |  |  |  |
| HbA1c (%) | 5.46 | 5.56 | -0.10 [-0.32, 0.10] | 0.306 |
| Total cholesterol (mmol/L) | 5.42 | 5.20 | 0.22 [-0.49, 0.93] | 0.522 |
| HDL-cholesterol (mmol/L) | 1.41 | 1.37 | 0.04 [-0.09, 0.19] | 0.480 |
| LDL-cholesterol (mmol/L) | 3.35 | 3.29 | 0.06 [-0.55, 0.68] | 0.828 |
| Triglycerides (mmol/L) | 1.47 | 1.19 | 0.28 [-0.26, 0.82] | 0.290 |
| **Dietary intake** |  |  |  |  |
| Total calories (kcal) | 1677.7 | 1324.4 | 353.3 [-84.00, 790.58] | 0.108 |
| Protein (g) | 75.12 | 54.91 | 20.21 [-3.23, 43.65] | 0.088 |
| Carbohydrates (g) | 193.33 | 164.87 | 28.47 [-12.38, 69.31] | 0.163 |
| Fat (g) | 65.41 | 51.08 | 14.33 [-12.24, 40.90] | 0.277 |

Abbreviations: HbA1c – glycated hemoglobin aka hemoglobin A1c; HDL- high-density lipoprotein; LDL – low-density lipoprotein
Values are adjusted means with 95% confidence intervals derived from analysis of covariance (ANCOVA), adjusting for baseline measurements. \**P* was obtained using ANCOVA, indicating between-group comparisons at each time point.

For blood parameters, a modest difference was observed in HbA1c levels, with the MyGeneMyDiet® group exhibiting slightly lower levels compared to the standard recommendation group at month 6 (5.49% vs. 5.60%; difference: 0.11 [95% CI: 0.01, 0.20], P = 0.038). However, there was insufficient evidence of differences in other blood parameters, along with total calorie and macronutrient intakes.

### Comparison of Outcomes Between Intervention Groups at Month 12 (Inactive Phase)

At the end of the inactive phase (month 12), the overall pattern observed at month 6 persisted for both anthropometric and biochemical markers, with no meaningful differences between the two groups (Table 2). Compared to the standard recommendation group, the MyGeneMyDiet® group reported lower intake of total calories (difference: 353.3 kcal [95% CI: −84.00, 790.58], P = 0.108), protein (difference 20.21 g ([95% CI: −3.23, 43.65], P = 0.088), carbohydrate (difference 28.47 g ([95% CI: −12.38, 69.31], P = 0.163), and fat (difference 14.33 g ([95% CI: −12.24, 40.90], P = 0.277).

### Within-Group Changes from Baseline

Compared to baseline, at month 6, both groups exhibited reductions in dietary intake, with the standard recommendation group showing a decrease in total calorie intake (−359.1 kcal, P = 0.030) and protein intake (−12.2 g, P = 0.042), while the MyGeneMyDiet® group had reductions in total calorie intake (−295.8 kcal, P = 0.015) and carbohydrate intake (−35.5 g, P = 0.010). Additionally, a modest reduction in waist circumference was observed in the standard recommendation group (−1.5 cm, P = 0.009).

Conversely, most anthropometric and biochemical markers showed little to no change in either group. Weight, BMI, body fat percentage, and lipid profile measures remained largely stable over the 6-month period. HbA1c levels were also unchanged in both groups (Table 3).

**Table 3.** Changes in outcome variables in month 6 relative to baseline.

|  | Standard Recommendation<br>(*n* = 23) |  |  |  | MyGeneMyDiet®<br>(*n* = 29) |  |  |  |
| --- | --- | --- | --- | --- | --- | --- | --- | --- |
|  | Baseline | Month 6 | Change | *P*-value* | Baseline | Month 6 | Change | *P*-value* |
| **Anthropometry** |  |  |  |  |  |  |  |  |
| Weight (kg) | 76.6 ± 14.2 | 75.9 ± 13.4 | -0.7 | 0.179 | 75.3 ± 13.5 | 74.9 ± 14.6 | -0.4 | 0.447 |
| BMI (kg/m<sup>2</sup>) | 29.0 ± 2.9 | 29.0 ± 3.0 | 0.0 | 0.742 | 29.5 ± 4.1 | 29.4 ± 4.4 | -0.5 | 0.352 |
| Waist circumference (cm) | 90.2 ± 9.2 | 88.7 ± 8.8 | -1.5 | 0.009 | 90.0 ± 10.5 | 88.8 ± 11.3 | -1.2 | 0.131 |
| Body fat (%) | 33.8 ± 7.6 | 33.6 ± 8.2 | -0.2 | 0.551 | 38.3 ± 6.8 | 38.1 ± 6.7 | -0.2 | 0.486 |
| **Blood parameters** |  |  |  |  |  |  |  |  |
| HbA1c (%) | 5.6 ± 0.4 | 5.6 ± 0.3 | 0.0 | 0.103 | 5.5 ± 0.4 | 5.5 ± 0.4 | 0.0 | 0.655 |
| Total cholesterol (mmol/L) | 5.5 ± 1.0 | 5.2 ± 1.0 | -0.3 | 0.246 | 5.4 ± 1.0 | 5.3 ± 1.0 | -0.1 | 0.294 |
| HDL-cholesterol (mmol/L) | 1.3 ± 0.3 | 1.3 ± 0.3 | 0.0 | 0.483 | 1.4 ± 0.3 | 1.3 ± 0.3 | -0.1 | 0.072 |
| LDL-cholesterol (mmol/L) | 3.5 ± 0.9 | 3.2 ± 0.9 | -0.3 | 0.180 | 3.4 ± 0.9 | 3.2 ± 1.0 | -0.2 | 0.087 |
| Triglycerides (mmol/L) | 1.6 ± 0.9 | 1.5 ± 0.6 | -0.1 | 0.773 | 1.4 ± 0.6 | 1.7 ± 1.7 | 0.3 | 0.341 |
| **Dietary intake** |  |  |  |  |  |  |  |  |
| Total calories (kcal) | 2036.2 ± 556.9 | 1677.1 ± 437.5 | -359.1 | 0.030 | 1811.7 ± 443.0 | 1515.9 ± 502.2 | -295.8 | 0.015 |
| Protein (g) | 79.5 ± 23.1 | 67.3 ± 18.4 | -12.2 | 0.042 | 68.7 ± 21.1 | 61.0 ± 20.5 | -7.7 | 0.085 |
| Carbohydrates (g) | 226.6 ± 89.0 | 196.1 ± 60.8 | -30.5 | 0.184 | 205.7 ± 54.8 | 170.2 ± 54.8 | -35.5 | 0.010 |
| Fat (g) | 86.1 ± 35.5 | 68.7 ± 27.9 | -17.4 | 0.058 | 75.7 ± 29.6 | 65.5 ± 27.7 | -10.2 | 0.126 |

Abbreviations: HbA1c – glycated hemoglobin aka hemoglobin A1c; HDL- high-density lipoprotein; LDL – low-density lipoprotein
Values are presented as mean ± SD; \**P* was obtained using paired *t*-test
The minus sign (-) indicates a reduction from baseline. All other change values indicate no change or an increase.

Meanwhile, when comparing the baseline and month 12 results, both groups exhibited reductions in dietary intake, with the MyGeneMyDiet® group reporting lower total calorie (−461.4 kcal, P = 0.001), protein (−12.1 g, P = 0.007), carbohydrate (−46.4 g, P = 0.015), and fat intake (−22.0 g, P = 0.014) compared to baseline. In the standard recommendation group, reductions in dietary intake were observed but were more modest and did not reach statistical significance.

For anthropometric measures, both groups showed slight reductions in weight and BMI, but these changes remained minimal over 12 months. A decrease in waist circumference was observed in the standard recommendation group (−3.2 cm, P = 0.010), while a similar trend in the MyGeneMyDiet® group (−3.0 cm, P = 0.117) did not reach statistical significance. Body fat percentage showed small reductions in both groups but remained largely unchanged from baseline. HbA1c, lipid profiles, and triglycerides remained stable in both groups, with no meaningful changes over time (Table 4).

**Table 4.** Changes in outcome variables in month 12 relative to baseline.

|  | Standard Recommendation<br>(*n* = 12) |  |  |  | MyGeneMyDiet®<br>(*n* = 15) |  |  |  |
| --- | --- | --- | --- | --- | --- | --- | --- | --- |
|  | Baseline | Month 12 | Change | *P*-value* | Baseline | Month 12 | Change | *P*-value* |
| **Anthropometry** |  |  |  |  |  |  |  |  |
| Weight (kg) | 72.9 ± 11.9 | 71.7 ± 11.4 | -1.2 | 0.227 | 72.9 ± 10.7 | 71.7 ± 13.3 | -1.2 | 0.343 |
| BMI (kg/m<sup>2</sup>) | 28.3 ± 1.5 | 27.9 ± 1.9 | -0.4 | 0.225 | 28.6 ± 3.0 | 28.0 ± 3.7 | -0.6 | 0.281 |
| Waist circumference (cm) | 87.8 ± 4.7 | 84.6 ± 4.6 | -3.2 | 0.010 | 87.4 ± 7.5 | 84.4 ± 9.3 | -3.0 | 0.117 |
| Body fat (%) | 33.7 ± 7.0 | 32.7 ± 7.2 | -1.0 | 0.055 | 37.6 ± 6.9 | 36.3 ± 7.3 | -1.0 | 0.154 |
| **Blood parameters** |  |  |  |  |  |  |  |  |
| HbA1c (%) | 5.6 ± 0.4 | 5.5 ± 0.4 | -0.1 | 0.412 | 5.5 ± 0.5 | 5.5 ± 0.6 | 0.0 | 0.546 |
| Total cholesterol (mmol/L) | 5.8 ± 1.0 | 5.5 ± 1.0 | -0.3 | 0.216 | 5.4 ± 1.0 | 5.1 ± 0.9 | -0.3 | 0.318 |
| HDL-cholesterol (mmol/L) | 1.3 ± 0.3 | 1.4 ± 0.3 | 0.1 | 0.487 | 1.4 ± 0.4 | 1.4 ± 0.3 | 0.0 | 0.487 |
| LDL-cholesterol (mmol/L) | 3.8 ± 0.9 | 3.5 ± 1.0 | -0.3 | 0.144 | 3.4 ± 0.9 | 3.2 ± 0.8 | -0.2 | 0.387 |
| Triglycerides (mmol/L) | 1.5 ± 0.5 | 1.5 ± 0.9 | 0.0 | 0.891 | 1.3 ± 0.5 | 1.2 ± 0.4 | -0.1 | 0.350 |
| **Dietary intake** |  |  |  |  |  |  |  |  |
| Total calories (kcal) | 2025.3 ± 716.8 | 1718.3 ± 620.2 | -307 | 0.236 | 1753.3 ± 359.3 | 1291.9 ± 466.5 | -461.4 | 0.001 |
| Protein (g) | 79.1 ± 26.9 | 79.8 ± 40.6 | 0.7 | 0.955 | 63.3 ± 18.5 | 51.2 ± 16.3 | -12.1 | 0.007 |
| Carbohydrates (g) | 239.8 ± 109.0 | 199.7 ± 46.4 | -40.1 | 0.128 | 206.2 ± 57.7 | 159.8 ± 63.7 | -46.4 | 0.015 |
| Fat (g) | 83.3 ± 39.0 | 67.0 ± 43.1 | -16.3 | 0.288 | 71.8 ± 27.5 | 49.8 ± 22.3 | -22.0 | 0.014 |

Abbreviations: HbA1c – glycated hemoglobin aka hemoglobin A1c; HDL- high-density lipoprotein; LDL – low-density lipoprotein
Values are presented as mean ± SD; \**P* was obtained using paired *t*-tests
The minus sign (-) indicates a reduction from baseline. All other change values indicate no change or an increase.

### Additional Analyses to Address Potential Issues on Attrition and Adherence

To address potential issues related to attrition and adherence, additional sensitivity analyses were conducted using Inverse Probability of Attrition Weighting (IPAW) and Last Observation Carried Forward (LOCF). These methods were applied to account for missing data, to assess the effect of the intervention in the presence of dropouts and incomplete follow-up. Results for the active and inactive phases using these methods are provided in Supplementary Tables 4 and 5. Results obtained with LOCF and IPAW yielded largely similar results as with the raw (unadjusted, complete case) analysis.

## Discussion

This study evaluated the utility of gene-based versus standard weight recommendations on anthropometric, specific biochemical markers (glucose and lipid profile), and dietary outcomes over 12 months. Although changes within each group were observed, we found no evidence to suggest differences between the MyGeneMyDiet® and standard recommendation groups across most outcomes at both the active (month 6) and inactive (month 12) phases.

No significant differences in anthropometric outcomes were observed between the groups throughout the trial. While some studies have demonstrated reductions in BMI, body weight, and body fat with diets tailored to specific genetic polymorphisms [39–41], our findings are consistent with studies that reported no added benefit from nutrigenetic-guided diets [32, 42]. This lack of advantage may reflect the multifactorial nature of weight regulation, where genetic predisposition is only one component of a complex system influenced by baseline metabolic health, psychosocial readiness for dietary changes, and adherence to interventions.

Previous studies examining macronutrient-specific gene-diet interactions have reported improvements in HDL-cholesterol, triglycerides, and inflammatory markers [43–45]. However, apart from the reduction in HbA1c level in the MyGeneMyDiet® group at month 6, we did not observe significant improvements in biochemical markers. This may be attributed to low adherence rates or the less targeted nature of our dietary and lifestyle recommendations.

Dietary intake patterns showed notable reductions, with MyGeneMyDiet® participants demonstrating lower caloric and macronutrient intake. These findings align with evidence suggesting that personalized recommendations, particularly when informed by genetic information, can positively influence dietary behaviors [22, 23, 32]. The perception of personalized genetic information may have enhanced participant engagement and accountability, potentially facilitating these changes [46]. However, the difference in dietary intake between groups may also be partly attributed to the higher proportion of female participants in the MyGeneMyDiet® group. Women tend to report lower energy intake and may have different dietary behaviors and responses to nutrition interventions compared to men [47–49]. While genetic-based recommendations may have influenced dietary choices, participant characteristics, including sex distribution, should also be considered when interpreting the observed changes. Despite these reductions in dietary intake, the changes were insufficient to produce meaningful differences in anthropometric or biochemical outcomes, underscoring the critical role of adherence and sustained behavioral support in achieving long-term weight management.

The MyGeneMyDiet® trial’s two-phase design provided unique insights into the immediate and long-term impacts of personalized interventions. Offering genetic counseling may have further enhanced participant engagement by providing a personalized context for the recommendations. This underscores the value of integrating education and support into personalized nutrition strategies.

To our knowledge, this is among the first studies to evaluate gene-based weight-loss interventions in a low- and middle-income country (LMIC) setting. Implementing nutrigenomics research in resource-limited environments poses unique challenges, as highlighted in our previous work [50]. These include limited genomic literacy among participants, infrastructural barriers, and sociocultural factors influencing dietary and lifestyle behaviors. Additionally, broader systemic challenges such as access to healthcare and nutrition education further complicate implementation. Tailoring interventions to these unique contexts is essential for the scalability and sustainability of personalized nutrition strategies in LMICs.

Adherence to dietary and physical activity recommendations is a key determinant of success in weight-loss interventions [51, 52]. Low adherence rates might have influenced our results, despite sensitivity analyses (IPAW and LOCF) confirming the modest differences observed. The COVID-19 pandemic, which coincided with the intervention period, introduced disruptions such as movement restrictions and shifts in daily routines, likely exacerbating adherence challenges. This highlights the importance of ongoing behavioral support to mitigate external disruptions.

Our findings suggest that both gene-based and standard recommendations can yield comparable outcomes, even in resource-limited settings. When contextualized within the broader literature, these findings raise questions about the added value of gene-based strategies in routine clinical practice. While gene-based recommendations may facilitate modest changes in dietary behavior, particularly macronutrient intake, they do not appear to outperform standard recommendations in achieving anthropometric or biochemical improvements [25, 27].

In conclusion, both gene-based and standard nutritional recommendations resulted in comparable outcomes in weight-loss interventions. Future research should focus on understanding the factors that influence adherence, optimizing the delivery of personalized recommendations, and evaluating their effectiveness in diverse real-world settings.

## Acknowledgements

We thank the administrative and infrastructure support of DOST-Food and Nutrition Research Institute, and the study participants for their commitment to the trial. We would also like to thank the members of the MyGeneMyDiet® Scientific Advisory Board, namely, Dr. Celeste C. Tanchoco, Dr. Liezl M. Atienza, Dr. Ma-am Joy R. Tumulak, Ms. Zenaida F. Velasco, Mr. Peter James B. Abad, Dr. Jan Paolo D. Dipasupil, and Ms. Virgith B. Buena. Likewise, the authors would like to acknowledge Rachel Evangelista for the operational support.

The authors’ responsibilities are as follows – JSN, JPHL: conceived and designed the study; JSN, JPHL, DGDR, MPR: development of overall research plan; JSN, JCR, GBG: analyzed the data; JSN, JPHL, DGDR, MPR, MLM, RDF, NLCS: study oversight; AMFDD, JJVC, MGF, DJVF, DASM, MVP, HSSM, RVMC, AAB, GMA, RCB: conducted research, collected data, assembled and organized the data; FJBD, GBG: guidance and oversight during the data analysis and manuscript preparation; JSN: wrote the paper and had primary responsibility for the final content. All authors critically reviewed the manuscript and approved of the final version.

## Data Availability

Data for this study is available upon request through a data transfer agreement with the Food and Nutrition Research Institute. Interested researchers may contact the corresponding author to initiate the request.

## Funding

This research was funded by the Department of Science and Technology – Food and Nutrition Research Institute through the Locally Funded Project (LFP) of the Philippines’ Department of Budget and Management.

## Conflict of interest

The authors declare no competing interest.

## Abbreviations

BMI: body mass index
DNA: deoxyribonucleic acid
FCT: Food Composition Table
FIERC: FNRI Institutional Ethics Review Committee
FNRI: Food and Nutrition Research Institute
FTO: fat mass and obesity-associated gene
HbA1c: glycated hemoglobin
HPLC: High-performance liquid chromatography
IPAQ-SF: International Physical Activity Questionnaire – Short Form
IPAW: Inverse Probability of Attrition Weighting
ITT: Intention-to-Treat analysis
LOCF: Last Observation Carried Forward
RNDs: Registered Nutritionist-Dietitians
SD: standard deviation
SNOSE: sequentially numbered, opaque sealed envelopes
TCF7L2: transcription factor 7-like 2 gene
UCP1: uncoupling protein 1 gene
WHO: World Health Organization

## Notes

### Competing Interest Statement

The authors have declared no competing interest.

### Clinical Trial

NCT05098899

### Funding Statement

The study was funded by the Department of Budget and Management, Philippines through the Locally Funded Project (LFP).

### Author Declarations

FNRI Institutional Ethics Review Committee (FIERC) of DOST-Food and Nutrition Research Institute gave ethical approval for this work.

## References

1. Lamiquiz-Moneo I, Mateo-Gallego R, Bea AM, Dehesa-García B, Pérez-Calahorra S, Marco-Benedí V, et al. Genetic predictors of weight loss in overweight and obese subjects. Sci Rep. 2019;9(1):10770, doi: 10.1038/s41598-019-47283-5.

2. Gkouskou KK, Grammatikopoulou MG, Lazou E, Vasilogiannakopoulou T, Sanoudou D, Eliopoulos AG. A genomics perspective of personalized prevention and management of obesity. Hum Genomics. 2024 Jan 29;18(1):4. doi: 10.1186/s40246-024-00570-3. PMID: 38281958; PMCID: PMC10823690.

3. Mera-Charria A, Nieto-Lopez F, Francès MP, Arbex PM, Vila-Vecilla L, Russo V, Silva CCV, De Souza GT. Genetic variant panel allows predicting both obesity risk, and efficacy of procedures and diet in weight loss. Front Nutr. 2023 Nov 16;10:1274662. doi: 10.3389/fnut.2023.1274662. PMID: 38035352; PMCID: PMC10687570.

4. Schlauch KA, Read RW, Lombardi VC, Elhanan G, Metcalf WJ, Slonim AD, et al. A comprehensive genome-wide and phenome-wide examination of BMI and obesity in a Northern Nevadan Cohort. G3 (Bethesda). 2020;10(2):645–64, doi: 10.1534/g3.119.400910.

5. Jiang L, Penney KL, Giovannucci E, Kraft P, Wilson KM. A genome-wide association study of energy intake and expenditure. PLoS One. 2018;13(8): e0201555, doi: 10.1371/journal.pone.0201555.

6. Dougkas A, Yaqoob P, Givens DI, Reynolds CK, Minihane AM. The impact of obesity-related SNP on appetite and energy intake. Br J Nutr. 2013;110(6):1151–6, doi: 10.1017/S0007114513000147.

7. Cho HW, Jin HS, Eom YB. The interaction between FTO rs9939609 and physical activity is associated with a 2-fold reduction in the risk of obesity in Korean population. Am J Hum Biol. 2021;33(3):e23489, doi: 10.1002/ajhb.23489.

8. Chermon D, Birk R. FTO common obesity SNPs interact with actionable environmental factors: physical activity, sugar-sweetened beverages and wine consumption. Nutrients. 2022;14(19), htpps://doi: 10.3390/nu14194202.

9. Rana S, Nawaz H. Interactive effects of FTO rs9939609 and obesogenic behavioral factors on adiposity-related anthropometric and metabolic phenotypes. Nucleosides Nucleotides Nucleic Acids. 2023;42(8):637–56, doi: 10.1080/15257770.2023.2182886.

10. Kilpeläinen TO, Qi L, Brage S, Sharp SJ, Sonestedt E, Demerath E, et al. Physical activity attenuates the influence of FTO variants on obesity risk: a meta-analysis of 218,166 adults and 19,268 children. PLoS Med. 2011;8(11):e1001116, doi: 10.1371/journal.pmed.1001116.

11. Chathoth S, Ismail MH, Vatte C, Cyrus C, Al Ali Z, Ahmed KA, et al. Association of Uncoupling Protein 1 (UCP1) gene polymorphism with obesity: a case-control study. BMC Med Genet. 2018;19(1):203, doi: 10.1186/s12881-018-0715-5.

12. Cannon B, Nedergaard J. Brown adipose tissue: function and physiological significance. Physiol Rev. 2004;84(1):277–359, doi: 10.1152/physrev.00015.2003.

13. Nagai N, Sakane N, Kotani K, Hamada T, Tsuzaki K, Moritani T. Uncoupling protein 1 gene −3826 A/G polymorphism is associated with weight loss on a short-term, controlled-energy diet in young women. Nutr Res. 2011;31(4):255–61, doi: 10.1016/j.nutres.2011.03.010.

14. Fumeron F, Durack-Bown I, Betoulle D, Cassard-Doulcier AM, Tuzet S, Bouillaud F, et al. Polymorphisms of uncoupling protein (UCP) and beta 3 adrenoreceptor genes in obese people submitted to a low calorie diet. Int J Obes Relat Metab Disord. 1996;20(12):1051–4.

15. Kogure A, Yoshida T, Sakane N, Umekawa T, Takakura Y, Kondo M. Synergic effect of polymorphisms in uncoupling protein 1 and beta3-adrenergic receptor genes on weight loss in obese Japanese. Diabetologia. 1998;41(11):1399, doi: 10.1007/s001250051084.

16. Grau K, Cauchi S, Holst C, Astrup A, Martinez JA, Saris WH, et al. TCF7L2 rs7903146-macronutrient interaction in obese individuals’ responses to a 10-wk randomized hypoenergetic diet. Am J Clin Nutr. 2010;91(2):472–9, doi: 10.3945/ajcn.2009.27947.

17. Bride L, Naslavsky M, Lopes Yamamoto G, Scliar M, Pimassoni LH, Sossai Aguiar P, et al. TCF7L2 rs7903146 polymorphism association with diabetes and obesity in an elderly cohort from Brazil. PeerJ. 2021;9:e11349, doi: 10.3945/ajcn.2009.27947.

18. Phillips CM, Goumidi L, Bertrais S, Field MR, McManus R, Hercberg S, et al. Dietary saturated fat, gender and genetic variation at the TCF7L2 locus predict the development of metabolic syndrome. J Nutr Biochem. 2012;23(3):239–44, doi: 10.1016/j.jnutbio.2010.11.020.

19. Muller YL, Hanson RL, Piaggi P, Chen P, Wiessner G, Okani C, et al. Assessing the role of 98 established loci for BMI in American Indians. Obesity (Silver Spring). 2019;27(5):845–54, doi: 10.1002/oby.22433.

20. Al-Daghri NM, Alkharfy KM, Al-Attas OS, Krishnaswamy S, Mohammed AK, Albagha OM, et al. Association between type 2 diabetes mellitus-related SNP variants and obesity traits in a Saudi population. Mol Biol Rep. 2014;41(3):1731–40, doi: 10.1007/s11033-014-3022-z.

21. Abadi A, Alyass A, Robiou du Pont S, Bolker B, Singh P, Mohan V, et al. Penetrance of polygenic obesity susceptibility loci across the body mass index distribution. Am J Hum Genet. 2017;101(6):925–38, doi: 10.1016/j.ajhg.2017.10.007.

22. Nielsen DE, El-Sohemy A. Disclosure of genetic information and change in dietary intake: a randomized controlled trial. PLoS One. 2014;9(11):e112665, doi: 10.1371/journal.pone.0112665.

23. Horne J, Gilliland J, O’Connor C, Seabrook J, Madill J. Enhanced long-term dietary change and adherence in a nutrigenomics-guided lifestyle intervention compared to a population-based (GLB/DPP) lifestyle intervention for weight management: results from the NOW randomised controlled trial. BMJ Nutr Prev Health. 2020;3(1):49–59, doi: 10.1136/bmjnph-2020-000073.

24. Celis-Morales C, Marsaux CF, Livingstone KM, Navas-Carretero S, San-Cristobal R, Fallaize R, et al. Can genetic-based advice help you lose weight? Findings from the Food4Me European randomized controlled trial. Am J Clin Nutr. 2017;105(5):1204–13, doi: 10.3945/ajcn.116.145680.

25. Coletta AM, Sanchez B, O’Connor A, Dalton R, Springer S, Koozehchian MS, et al. Alignment of diet prescription to genotype does not promote greater weight loss success in women with obesity participating in an exercise and weight loss program. Obes Sci Pract. 2018;4(6):554–74, doi: 10.1002/osp4.305.

26. Doets EL, de Hoogh IM, Holthuysen N, Wopereis S, Verain MCD, van den Puttelaar J, et al. Beneficial effect of personalized lifestyle advice compared to generic advice on wellbeing among Dutch seniors - An explorative study. Physiol Behav. 2019; 210:112642, doi: 10.1016/j.physbeh.2019.112642.

27. Marsaux CF, Celis-Morales C, Livingstone KM, Fallaize R, Kolossa S, Hallmann J, et al. Changes in physical activity following a genetic-based internet-delivered personalized intervention: randomized controlled trial (Food4Me). J Med Internet Res. 2016;18(2):e30, doi: 10.2196/jmir.5198.

28. O’Donovan CB, Walsh MC, Forster H, Woolhead C, Celis-Morales C, Fallaize R, et al. The impact of MTHFR 677C → T risk knowledge on changes in folate intake: findings from the Food4Me study. Genes Nutr. 2016;11:25, doi: 10.1186/s12263-016-0539-x.

29. ElGendy K, Malcomson FC, Lara JG, Bradburn DM, Mather JC. Effects of dietary interventions on DNA methylation in adult humans: systematic review and meta-analysis. Br J Nutr. 2018; 120(9):961–976.doi:10.1017/S000711451800243X.

30. Brennan L, de Roos B. Nutrigenomics: lessons learned and future perspectives. Am J Clin Nutr, 2021; 113(3):503–516.doi:10.1093/ajcn/nqaa366.

31. Aljasir S, Eid NMS, Volpi EV, Tewfik I. Nutrigenomics-guided lifestyle intervention programmes: A critical scoping review with directions for future research. Clinical Nutrition ESPEN. 2024; 64:296–306. doi10.1016/j.clnesp.2024.10.149.

32. Aldubayan MA, Pigborg K, Gormsen SMO, et al. A double-blind, randomized parallel intervention to evaluate biomarker-based nutrition plans for weight loss: the PREVENTOMICS study. Clin Nutr. 2022; 41(8):1834–1844. doi:10.1016/j.clnu.2022.06.032.

33. Meisel SF, Beeken RJ, van Jaarsveld CH, Wardle J. Genetic susceptibility testing and readiness to control weight: Results from a randomized controlled trial. Obesity (Silver Spring). 2015;23(2):305–312.doi:10.1002/oby.20958.

34. Tan PY, Moore JB, Bai L, Tang G, Gong YY. In the context of the triple burden of malnutrition: A systematic review of gene-diet interactions and nutritional status. Crit Rev Food Sci Nutr. 2024; 64(11):3235–3263.doi:10.1080/10408398.2022.2131727.

35. Martinez-Lopez E, Garcia-Garcia MR, Gonzales-Avalos JM, et al. Effect of Ala54Thr polymorphism on FABP2 on anthropometric and biochemical variables in response to moderate-fat diet. Nutrition. 2013;29(1):46–51.doi:10.1016/j.nut.2012.03.002

36. Griffin BA, Walker CG, Jebb SA, et al. APOE4 genotype exerts greater benefit in lowering plasma cholesterol and apolipoprotein B than wild type (E3/E3), after replacement of dietary saturated fats with low glycemic index carbohydrates. Nutrients. 2018;10(10):1524.doi:10.3390/nu10101524.

37. Nacis JS, Labrador JPH, Ronquillo DGD, Rodriguez MP, Dablo AMFD, Frane RD, et al. A study protocol for a pilot randomized controlled trial to evaluate the effectivess of a gene-based nutrition and lifestyle recommendation for weight management among adults: the MyGeneMyDiet® study. Front. Nutr. 2023;10.1228234. doi:10.3389/fnut.2023.1238234.

38. Gallagher D, Heymsfield SB, Heo M, Jebb SA, Murgatroyd PR, Sakamoto Y. Healthy percentage body fat ranges: an approach for developing guidelines based on body mass index. Am J Clin Nutr. 2000;72(3):694–701. doi:10.1093/ajcn/72.3.694.

39. Arkadianos I, Valdes AM, Marinos E, Florou A, Gill RD, Grimaldi KA. Improved weight management using genetic information to personalize a calorie controlled diet. Nutr J. 2007;6:29. doi:10.1186/1475-2891-6-29.

40. Horne JR, Gilliland JA, O’Connor CP, Seabrook JA, Madill J. Change in weight, BMI, and body composition in a population-based intervention versus genetic-based intervention: the NOW Trial. Obesity (Silver Spring). 2020;28(8):1419–27. doi:10.1186/1475-2891-6-29.

41. Pérez-Beltrán YE, González-Becerra K, Rivera-Iñiguez I, Martínez-López E, Ramos-Lopez O, Alcaraz-Mejía M, et al. A nutrigenetic strategy for reducing blood lipids and low-grade inflammation in adults with obesity and overweight. Nutrients. 2023;15(20). doi:10.3390/nu15204324.

42. Frankwich KA, Egnatios J, Kenyon ML, Rutledge TR, Liao PS, Gupta S, et al. Differences in weight loss between persons on standard balanced vs nutrigenetic diets in a randomized controlled trial. Clin Gastroenterol Hepatol. 2015;13(9):1625–32.e1. doi:10.1016/j.cgh.2015.02.044.

43. Garcia-Rios A, Alcala-Diaz JF, Gomez-Delgado F, et al. Beneficial effect of CETP gene polymorphism in combination with a Mediterranean diet influencing lipid metabolism in metabolic syndrome patients: CORDIOPREV study. Clin Nutr. 2018; 37(1):229–234.doi:10.1016/j.clnu.2016.12.011

44. Pérez-Beltrán YE, González-Becerra K, Rivera-Iñiguez I, et al. A Nutrigenetic Strategy for Reducing Blood Lipids and Low-Grade Inflammation in Adults with Obesity and Overweight. Nutrients. 2023;15(20):4324. Published 2023 Oct 10. doi:10.3390/nu15204324.

45. De Luis D, Izaola O, Primo D, Aller R. Role of rs670 variant of APOA1 gene on metabolic response after a high fat vs a low fat hypocaloric diets in obese human subjects. J Diabetes Complications. 2019; 33(3):249–254.doi:10.1016/j.diacomp.2018.10.015.

46. Hietaranta-Luoma H-L, Tahvonen R, Iso-Touru T, Puolijoki H, Hopia A. An intervention study of individual, apoE genotype-based dietary and physical activity advice: impact on health behavior. J Nutrigenet Nutrigenomics. 2015;7:161–74, doi:10.1159/000371743.

47. Leblanc V, Bégin C, Corneau L, Dodin S, Lemieux S. Gender differences in dietary intakes: what is the contribution of motivational variables?. J Hum Nutr Diet. 2015;28(1):37–46. doi:10.1111/jhn.12213.

48. Bärebring L, Palmqvist M, Winkvist A, Augustin H. Gender differences in perceived food healthiness and food avoidance in a Swedish population-based survey: a cross sectional study. Nutr J. 2020;19(1):140. Published 2020 Dec 29. doi:10.1186/s12937-020-00659-0

49. Feraco A, Armani A, Amoah I, et al. Assessing gender differences in food preferences and physical activity: a population-based survey. Front Nutr. 2024;11:1348456. Published 2024 Feb 20. doi:10.3389/fnut.2024.1348456.

50. Nacis JS, Kamande P, Toni AT, et al. Barriers and enablers to the effective implementation of omics research in low- and middle-income countries. Nat Biotechnol. 2024;42(6):988–991.doi:10.1038/s41587-024-02274-4.

51. Wang D, Benito PJ, Rubio-Arias JA, Ramos-Campo DJ, Rojo-Tirado MA. Exploring factors of adherence to weight loss interventions in population with overweight/obesity: an umbrella review. Obes Rev 2024; 25(9):e13783. doi:10.1111/obr.13783.

52. Balfour J, Boster J. Physical Activity and Weight Loss Maintenance. In: StatPearls. Treasure Island (FL): StatPearls Publishing; June 20, 2023.

