## Supplementary Materials for "Effect of Gene-based vs. Standard Weight-Loss Recommendations on Anthropometry, Lipid and Glucose Markers, and Dietary Intake: The MyGeneMyDiet® Study"

**Supplementary Table 1. *Eligibility Criteria***

*Study Inclusion Criteria:*

- Overweight/obese
- 19-59 years old
- BMI equal or greater than 25 but not more than 40 kg/m^2^
- Normal to pre-disease state level of the following blood parameters:
  - Fasting blood glucose (4.11 – 6.05 mmol/L)
  - Lipid profile
    - Total cholesterol:
      - Desirable (<5.2 mmol/L)
      - Borderline high (5.2 – 6.2 mmol/L)
      - High (>6.2 mmol/L)
    - Triglycerides (<2.26 mmol/L)
    - HDL- cholesterol (>1.68 mmol/L)
    - VLDL (0.26 – 1.04 mmol/L)
    - LDL (0.3 – 3.37 mmol/L)
  - Cortisol (171 – 536 mmol/L)
  - Thyroid stimulating hormone (TSH) (0.270 – 4.200 nmol/L)
  - Triiodothyronine (T3) (1.3 – 3.1 nmol/L)
  - Thyroxine (T4) (66 – 181 nmol/L)
- Blood pressure (systolic: less than 120 mm Hg; diastolic: less than 80 mm Hg)
- Carrier of at least one of the risk alleles of the three target genetic polymorphisms:
  - A allele for FTO rs9939609
  - T allele for *TCF7L2* rs7903146
  - G allele for *UCP1* rs1800592

*Study Exclusion Criteria:*

- Individuals with elevated levels of fasting blood glucose, lipids, blood pressure, thyroid hormones, and cortisol
- Self-reported history of heart disease
- Participation in weight-loss program
- Adherence to a restrictive/therapeutic diet in the past 3 months
- Self-reported weight changes (greater than 3.0 kg)
- Planned or recent bariatric surgery
- Consumption of weight-altering medications and/or nutritional supplements that provide weight gain/loss in the past 6 months
- Clinical diagnosis of any mental disorder
- Current use of mental health medications
- (For females) – pregnant, nursing, or with self-declared intention to become pregnant during the study
- Current and anticipated enrollment in another research study

**Supplementary: Development of the MyGeneMyDiet® Recommendations**

The development of the MyGeneMyDiet® Recommendations followed three investigative phases: Explore, Develop, and Integrate (**Figure 1**). Each phase built upon the findings of the previous, culminating in the implementation of genotype-based nutritional guidance through a randomized controlled trial.

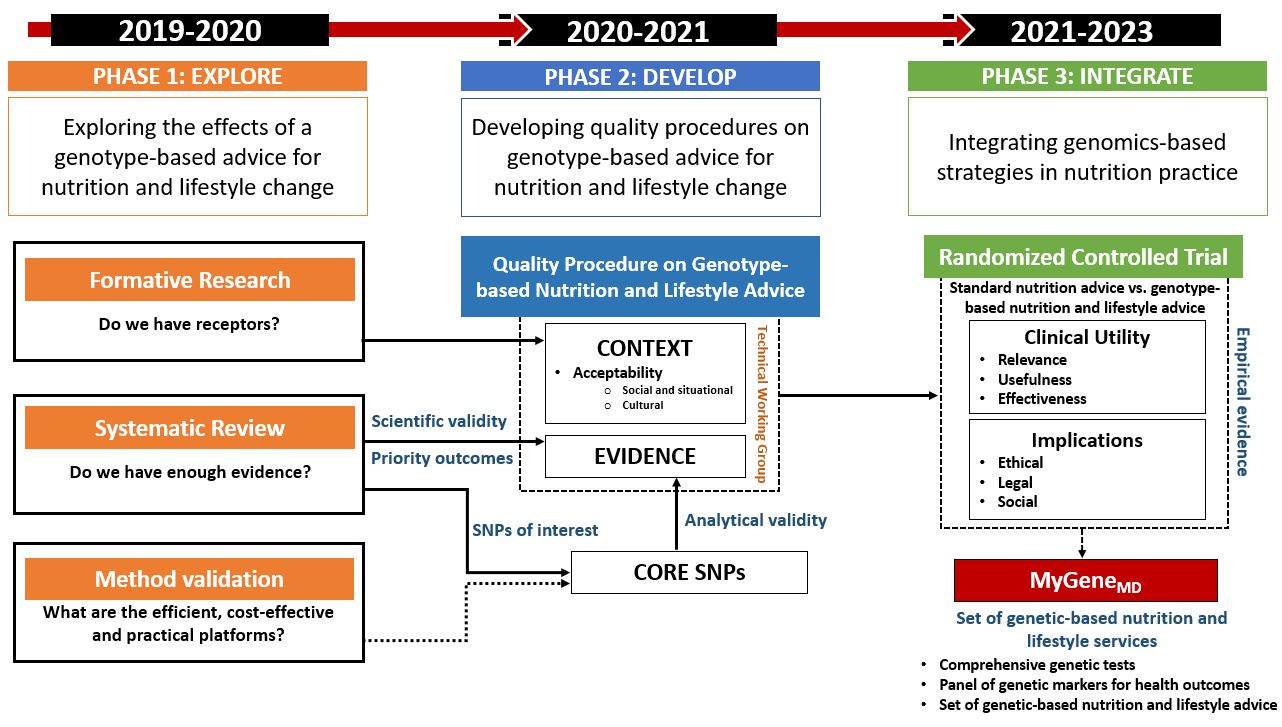

**Figure 1.** Development Stages of the MyGeneMyDiet® Recommendations

**Phase I: Explore**

This phase focused on:

1. Conducting a scoping review to assess methodologies used in genomics-based nutrition interventions.
2. Benchmarking of existing nutrigenetic tests.
3. Performing formative qualitative research to understand perceptions of nutrigenomics and gene-based nutrition recommendations among prospective end-users.
4. Validating laboratory procedures for genetic analysis.

*Identification of the Single Nucleotide Polymorphisms (SNPs) and the Phenotypes*

The scoping review (results not published) identified single nucleotide polymorphisms (SNPs) linked to obesity-related phenotypes. This review examined genetic testing, risk disclosure, and genotype-informed nutrition advice in randomized controlled trials (RCTs) targeting weight management. A total of 14 studies were identified as relevant (Figure 2).

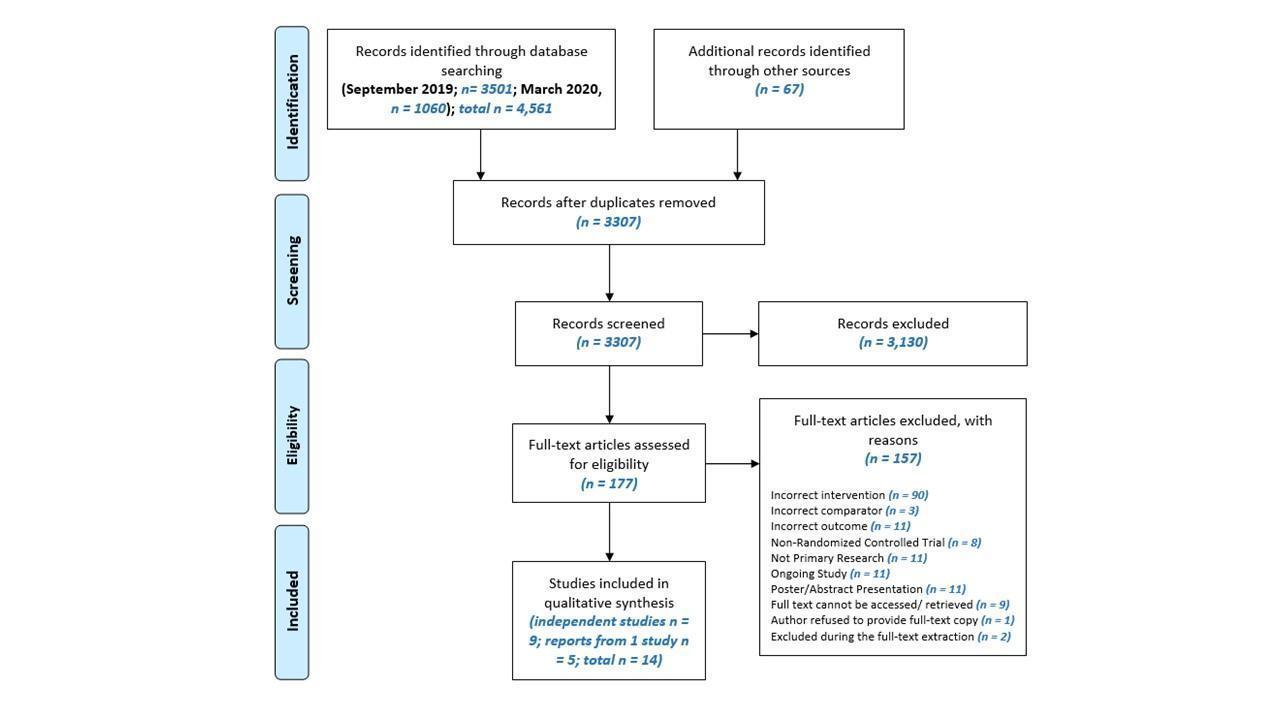

**Figure 2.** The flow of search, screening, and selection of studies following the PRISMA Diagram

Electronic searches were conducted in databases such as PubMed, Cochrane Library, EBSCO, MedLine, CINAHL, and Google Scholar using keywords (below) related to nutrigenomics, SNPs, precision nutrition, and weight management. Benchmarking was also conducted using sample test results and recommendations from nutrigenetic services.

*adult, nutrigenomics, nutritional genomics, nutrigenetics, nutritional genetics, personalized nutrition, nutritional epigenetics, single nucleotide polymorphisms, SNP, DNA, gene, gene*, genome*, precision nutrition, nutrition counsel*, nutrition advice, nutrition recommendation, dietary counsel*, dietary advice, dietary recommendation, genotype-based, gene-based, DNA-based, genomics-based, genetic test, genetic information, genetic risk, diet, food, nutrition, physical activity, smoking, alcohol.*

The pre-selected outcomes, assessed by at least two studies, included BMI, waist circumference, dietary fat intake, energy intake, and physical activity. The SNPs listed in Table 1 were identified for further development:

**Table 1.** Single nucleotide polymorphisms (SNPs) and phenotypes

| **Gene** | **SNP** | **Risk Allele/Variant** | **Phenotype** |
| --- | --- | --- | --- |
| *FTO* | rs99396609 | A (AA/AT) | BMI, waist circumference, physical activity levels |
| *TCF7L2* | rs7903146 | T (TT/CT) | Dietary fat intake |
| *UCP1** | rs1800592 | G (GG/GA) | Energy requirement |

******Included based on benchmarking of existing nutrigenetic recommendations*

**Phase II: Develop**

*Creation of Recommendation and Tools*

This phase concentrated on designing MyGeneMyDiet® Recommendations and their supporting tools. A Technical Working Group (TWG), consisting of nutritionist-dietitians and medical technologists from the Department of Science and Technology – Food and Nutrition Research Institute (DOST-FNRI), led the development process. Recommendations were based on:

- Findings from the scoping review.
- Evidence-based guidelines for managing obesity.
- Culturally appropriate content for local use.

The following tools and procedures were developed:

| **MyGeneMyDiet® standard tools, and quality standard procedures** | **Purpose/Contents** |
| --- | --- |
| Summary of Gene-based Nutrition Recommendations | Gene-based Nutrition Recommendations for risk allele carriers of *FTO* rs9939609, *UCP1* rs1800592, and *TCF7L2* rs7903146 |
| Standard Procedures for Gene-based Nutrition Assessment and Counseling | Process, procedures, and key persons involved in the gene-based nutrition assessment and counseling |
| Decision Trees of Recommendations | Comprises of algorithms of nutrition assessment procedures and corresponding MyGeneMyDiet® recommendations for *FTO* rs9939609, *UCP1* rs1800592, and *TCF7L2* rs7903146 |
| Nutrigenetic Report and Personalized Meal plans | A report detailing genetic test results, MyGeneMyDiet® recommendations, and sample meal plans |
| Nutrition Education Modules | A collection of nutritional education lessons focused on weight management strategies. Address topics related to healthy eating, physical activity, sustaining weight loss goals, and avoiding weight relapse |
| Genetic Education Flipcharts | A set of genetic education flipcharts designed for use by genetic counselors to communicate genetic test results and provide psychosocial counseling. |

*Scientific Advisory Board*

The Scientific Advisory Board (SAB), composed of nutritionist-dietitians, genetic counselors, and a lifestyle medicine physician, evaluated the recommendations and tools. Over five meeting held between February and December 2021, the SAB provided feedback, ensuring that the tools met scientific and professional standards (Table 2). Final recommendations were based on SAB consensus and iterative revisions.

**Table 2.** Summary of SAB meetings

| **SAB Meetings** | **Tools and Materials Reviewed** | **Date** |
| --- | --- | --- |
| 1^st^ SAB Meeting | Overview of the MyGeneMyDiet® Recommendations and Standard Procedures for Gene-based Nutrition Assessment | February 18, 2021 |
| 2^nd^ SAB Meeting | Nutrigenetic Report | March 18, 2021 |
| 3^rd^ SAB Meeting | Decision Trees for Recommendations | May 25, 2021 |
| 4^th^ SAB Meeting | Genetic Education Flipcharts | September 16, 2021 |
| 5^th^ SAB Meeting | Nutrition Education Modules | December 9, 2021 |

**Phase III: Integrate**

*Implementation and validation*

This phase tested the MyGeneMyDiet® Recommendations in a proof-of-concept randomized controlled trial (the MyGeneMyDiet® study). This phase provided empirical evidence for the relevance, utility, effectiveness of genotype-based nutrition advice.

*Final recommendations*

- *FTO* rs9939609 (risk allele A): Increased duration of moderate-to-vigorous physical activity, along with goals to achieve normal body weight and BMI.
- *UCP1* rs1800592 (risk allele G): An additional calorie deficit of up to 150 kcal from calculated energy needs.
- *TCF7L2* rs7903146 (risk allele T): Consumption of 15-20% of total energy from dietary fat.

Decision trees were developed to guide clinical decisions. Illustrated in Figure 3 is the sample decision tree for FTO

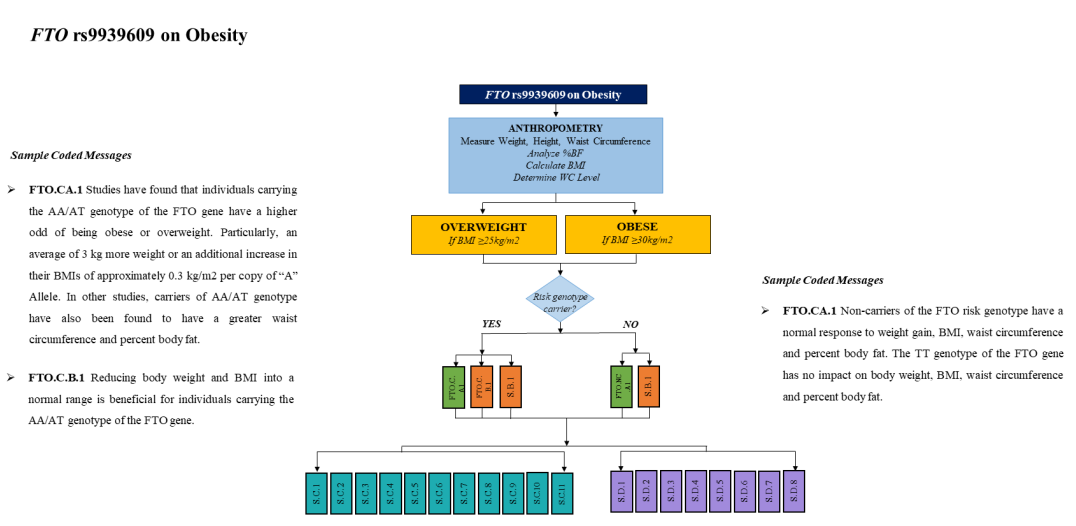

**Figure 3**. Sample decision tree for *FTO* rs9939609 on obesity

The decision trees are composed of four personalization domains:

- Domain A pertains to the disclosure of genotype and genetic risk information. These statements reiterate genetic test results and their relevance to nutrition and weight management.
- Domain B comprises the nutrition recommendations tailored fit on the results of the genetic test.
- Domain C encompasses dietary recommendations based on an individual's nutritional requirements.
- Domain D include recommendations for behaviors and lifestyles related to appropriate eating practices, physical activity, and the enhancement of general health and well-being.

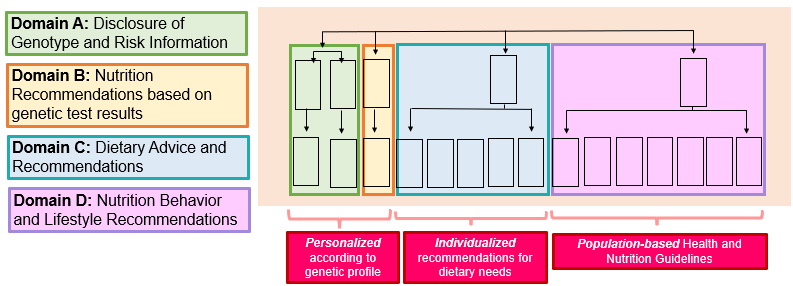

**Figure 4.** Personalization domains of the MyGeneMyDiet® Recommendations

The finalized tools and recommendations were detailed in supplementary materials in an earlier publication of the authors (Nacis JS, Labrador JPH, Ronquillo DGD, Rodriguez MP, Dablo AMFD, Frane RD, Madrid ML, Santos NLC, Carrillo JJV, Fernandez MG and Gonzales GBL (2023) A study protocol for a pilot randomized controlled trial to evaluate the effectiveness of a gene-based nutrition and lifestyle recommendation for weight management among adults: the MyGeneMyDiet® study. *Front. Nutr*. 10:1238234. doi: 10.3389/fnut.2023.1238234).

**Supplementary Table 2.** Distribution of genetic polymorphisms carried by the study participants.

| **Gene, rs number** |  | **Genotype distribution, n (%)^1^** | | | |
| --- | --- | --- | --- | --- | --- |
|  |  | **Active phase** | | **Inactive phase** | |
|  |  | **Standard Recommendation (*n* = 23)** | **MyGeneMyDiet® (*n* = 29)** | **Standard Recommendation (*n* = 12)** | **MyGeneMyDiet® (*n* = 15)** |
| *FTO,* rs9939609 | AA | 0 (0.0) | 3 (10.3) | 0 (0.0) | 1 (6.7) |
|  | AT | 11 (47.8) | 8 (27.6) | 8 (66.7) | 5 (33.3) |
| *UCP1,* rs1800592 | GG | 4 (17.4) | 6 (20.7) | 2 (16.7) | 3 (20.0) |
|  | GA | 12 (52.2) | 18 (62.1) | 4 (33.3) | 9 (60.0) |
| *TCF7L2,* rs7903146 | CC | 21 (91.3) | 29 (100.0) | 10 (83.3) | 15 (100.0) |
|  | CT | 2 (8.7) | 0 (0.0) | 2 (16.7) | 0 (0.0) |

^1^The genotypes presented here represent those carrying the risk alleles. Participants carrying the homozygous non-risk genotypes

(TT for *FTO* rs9939609, AA for *UCP1* rs1800592, and TT for *TCF7L2* rs7903146) were excluded from the study

**Supplementary Table 3.** Differences between groups for adherence with the recommendations at months 3, 6, 9, and 12 months ^1^

| **Time Point (Months)** | **TOTAL CALORIES** | | | **FAT INTAKE** | | | **PHYSICAL ACTIVITY** | | |
| --- | --- | --- | --- | --- | --- | --- | --- | --- | --- |
|  | **Standard**  **Recommendation** | **MyGeneMyDiet®** | ***P*-value** | **Standard**  **Recommendation** | **MyGeneMyDiet®** | ***P*-value** | **Standard**  **Recommendation** | **MyGeneMyDiet®** | ***P*-value** |
| 3 | 5 (21.73) | 3 (11.54) | 0.564 | 3 (13.04) | 5 (19.23) | 0.843 | 12 (63.16) | 8 (40.00) | 0.795 |
| 6 | 3 (13.04) | 7 (26.92) | 0.396 | 4 (17.39) | 6 (23.08) | 0.890 | 9 (56.25) | 7 (53.33) | 0.674 |
| 9 | 2 (11.76) | 4 (23.53) | 0.653 | 2 (11.76) | 4 (23.53) | 0.653 | 10 (66.67) | 3 (25.00) | 0.266 |
| 12 | 4 (33.33) | 2 (13.33) | 0.438 | 1 (8.33) | 2 (13.33) | 1.000 | 6 (66.67) | 3 (50.00) | 0.632 |

Values are reported as n (%); ^1^ Number of participants who adhered to the recommendations, as assessed through submitted 3-day food diaries and responses to the IPAQ-SF questionnaires.

**Supplementary Table 4.** Difference in outcomes at month 6 of intervention, with imputation methods

|  | **ACTIVE PHASE (Month 6)** | | | |
| --- | --- | --- | --- | --- |
|  | **Standard Recommendation** | **MyGeneMyDiet®** | **Difference [95% CI]** | ***P* – value*** |
| **Anthropometry** | | | | |
| Weight (kg) | 75.87 ± 13.48 | 74.92 ± 14.62 | RAW: 0.95 [0.89, 1.09]  LOCF: 0.63 [0.91, 1.11]  IPAW: 0.95 [0.89, 1.09] | 0.754  0.940  0.763 |
| BMI (kg/m^2^) | 28.96 ± 3.04 | 29.35 ± 4.37 | RAW: -0.39 [0.94, 1.08]  LOCF: -0.82 [0.95, 1.10]  IPAW: -0.39 [0.95, 1.09] | 0.813  0.571  0.682 |
| Waist circumference (cm) | 88.70 ± 8.85 | 88.81 ± 11.27 | RAW: -0.11 [0.94, 1.06]  LOCF: -1.23 [0.95, 1.07]  IPAW: -0.11 [0.94, 1.06] | 0.957  0.739  0.971 |
| Body fat (%) | 33.64 ± 8.24 | 38.12 ± 6.68 | RAW: -4.48 [1.02, 1.30]  LOCF: -4.44 [1.01, 1.30]  IPAW: -4.48 [1.04, 1.34] | 0.026  0.038  0.014 |
| **Biochemical markers** | | | | |
| HbA1c (%) | 5.64 ± 0.33 | 5.46 ± 0.37 | RAW: 0.18 [0.93, 1.00]  LOCF: 0.13 [0.94, 1.01]  IPAW: 0.18 [0.94, 1.01] | 0.063  0.173  0.212 |
| Total cholesterol (mmol/L) | 5.20 ± 1.01 | 5.28 ± 1.00 | RAW: -0.08 [0.91, 1.14]  LOCF: -0.08 [0.91, 1.14]  IPAW: -0.08 [0.91, 1.16] | 0.743  0.728  0.644 |
| HDL-cholesterol (mmol/L) | 1.29 ± 0.31 | 1.31 ± 0.29 | RAW: -0.02 [0.89, 1.16]  LOCF: -0.02 [0.90, 1.16]  IPAW: -0.02 [0.89, 1.17] | 0.843  0.782  0.783 |
| LDL-cholesterol (mmol/L) | 3.23 ± 0.95 | 3.19 ± 1.02 | RAW: 0.04 [0.81, 1.19]  LOCF: 0.01 [0.83, 1.20]  IPAW: 0.04 [0.82, 1.22] | 0.881  0.969  0.985 |
| Triglycerides (mmol/L) | 1.53 ± 0.66 | 1.71 ± 1.66 | RAW: -0.18 [0.74, 1.36]  LOCF: -0.13 [0.74, 1.31]  IPAW: -0.18 [0.75, 1.39] | 0.974  0.904  0.900 |
| **Dietary intake** | | | | |
| Total calories (kcal) | 1677.13 ± 437.5 | 1515.85 ± 502.2 | RAW: 161.28 [0.74, 1.05]  LOCF: 98.89 [0.77, 1.09]  IPAW: 161.28 [0.76, 1.08] | 0.157  0.312  0.254 |
| Protein (g) | 67.30 ± 18.39 | 61.0 ± 20.45 | RAW: 6.30 [0.74, 1.06]  LOCF: 2.13 [0.77, 1.12]  IPAW: 6.30 [0.74, 1.06] | 0.176  0.448  0.170 |
| Carbohydrates (g) | 196.15 ± 60.74 | 170.23 ± 54.81 | RAW: 25.92 [0.72, 1.05]  LOCF: 21.77 [0.73, 1.07]  IPAW: 25.92 [0.73, 1.07] | 0.146  0.208  0.209 |
| Fat (g) | 68.65 ± 27.88 | 65.54 ± 27.69 | RAW: 3.11 [0.74, 1.21]  LOCF: 0.14 [0.77, 1.26]  IPAW: 3.11 [0.75, 1.23] | 0.659  0.916  0.750 |

Abbreviations: HbA1c – glycated hemoglobin aka hemoglobin A1c; HDL- high-density lipoprotein; LDL – low-density lipoprotein

Values are presented as mean ± SD; **P* was obtained using *t*-tests

RAW indicates results without imputation; LOCF – Last Observation Carried Forward; IPAW – Inverse Probability of Attrition Weighting

**Supplementary Table 5.** Difference in outcomes at month 12 of intervention, with imputation methods

|  | **INACTIVE PHASE (Month 12)** | | | |
| --- | --- | --- | --- | --- |
|  | **Standard Recommendation** | **MyGeneMyDiet®** | **Difference [95% CI]** | ***P* – value*** |
| **Anthropometry** | | | | |
| Weight (kg) | 71.69 ± 11.4 | 71.68 ± 13.3 | RAW: 0.01 [0.87, 1.14]  LOCF: -0.67 [0.90, 1.11]  IPAW: 0.01 [0.84, 1.10] | 0.955  0.925  0.584 |
| BMI (kg/m^2^) | 27.86 ± 1.86 | 28.01 ± 3.71 | RAW: -0.15 [0.92, 1.08]  LOCF: -0.87 [0.95, 1.10]  IPAW: -0.15 [0.93, 1.09] | 0.995  0.563  0.858 |
| Waist circumference (cm) | 84.64 ± 4.61 | 84.42 ± 9.29 | RAW: 0.22 [0.93, 1.06]  LOCF: -2.07 [0.95, 1.09]  IPAW: 0.22 [0.93, 1.06] | 0.844  0.564  0.866 |
| Body fat (%) | 32.74 ± 7.19 | 36.3 ± 7.30 | RAW: -3.56 [0.94, 1.31]  LOCF: -4.54 [1.01, 1.30]  IPAW: -3.56 [0.99, 1.39] | 0.212  0.039  0.061 |
| **Biochemical markers** | | | | |
| HbA1c (%) | 5.48 ± 0.39 | 5.54 ± 0.59 | RAW: -0.06 [0.94, 1.08]  LOCF: 0.01 [0.96, 1.04]  IPAW: -0.06 [0.95, 1.12] | 0.815  0.915  0.433 |
| Total cholesterol (mmol/L) | 5.52 ± 1.07 | 5.13 ± 0.87 | RAW: 0.39 [0.81, 1.07]  LOCF: 0.15 [0.88, 1.07]  IPAW: 0.39 [0.80, 1.04] | 0.343  0.567  0.143 |
| HDL-cholesterol (mmol/L) | 1.37 ± 0.29 | 1.40 ± 0.33 | RAW: -0.04 [0.86, 1.22]  LOCF: -0.05 [0.92,1.18]  IPAW: -0.04 [0.85, 1.21] | 0.800  0.539  0.881 |
| LDL-cholesterol (mmol/L) | 3.47 ± 0.98 | 3.19 ± 0.78 | RAW: 0.27 [0.76, 1.16]  LOCF: 0.22 [0.78, 1.09]  IPAW: 0.27 [0.73, 1.08] | 0.553  0.361  0.221 |
| Triglycerides (mmol/L) | 1.50 ± 0.87 | 1.16 ± 0.41 | RAW: 0.34 [0.60, 1.10]  LOCF: -0.03 [0.70, 1.26]  IPAW: 0.34 [0.64, 1.15] | 0.179  0.692  0.299 |
| **Dietary intake** | | | | |
| Total calories (kcal) | 1718.3 ± 620.2 | 1291.9 ± 466.5 | RAW: 426.40 [0.56, 0.96]  LOCF: 184.37 [0.72, 1.04]  IPAW: 426.40 [0.49, 0.90] | 0.023  0.130  0.011 |
| Protein (g) | 79.75 ± 40.60 | 51.2 ± 16.31 | RAW: 28.55 [0.51, 0.87]  LOCF: 11.51 [0.69, 1.03]  IPAW: 28.55 [0.45, 0.88] | 0.003  0.092  0.009 |
| Carbohydrates (g) | 199.67 ± 46.43 | 159.8 ± 63.67 | RAW: 39.87 [0.59, 0.98]  LOCF: 22.43 [0.70, 1.04]  IPAW: 39.87 [0.58, 0.94] | 0.036  0.109  0.016 |
| Fat (g) | 67.0 ± 43.10 | 49.8 ± 22.32 | RAW: 17.20 [0.54, 1.11]  LOCF: 9.65 [0.68, 1.12]  IPAW: 17.20 [0.48, 1.10] | 0.160  0.280  0.124 |

Abbreviations: HbA1c – glycated hemoglobin aka hemoglobin A1c; HDL- high-density lipoprotein; LDL – low-density lipoprotein

Values are presented as mean ± SD; **P* was obtained using *t*-tests

RAW indicates results without imputation; LOCF – Last Observation Carried Forward; IPAW – Inverse Probability of Attrition Weighting
